# Identification of natural SARS-CoV-2 infection in seroprevalence studies among vaccinated populations

**DOI:** 10.1101/2021.04.12.21255330

**Authors:** Ryan T. Demmer, Brett Baumgartner, Talia D. Wiggen, Angela K. Ulrich, Ali J. Strickland, Brianna M. Naumchik, Bruno Bohn, Sara Walsh, Stephen Smith, Susan Kline, Steve D. Stovitz, Stephanie Yendell, Tim Beebe, Craig Hedberg

## Abstract

**Importance:** Identification of SARS-CoV-2 infection via antibody assays is important for monitoring natural infection rates. Most antibody assays cannot distinguish natural infection from vaccination.

**Objective:** To assess the accuracy of a nucleocapsid-containing assay in identifying natural infection among vaccinated individuals.

**Design:** A longitudinal cohort comprised of healthcare workers (HCW) in the Minneapolis/St. Paul metropolitan area was enrolled. Two rounds of seroprevalence studies separated by one month were conducted from 11/2020–1/2021. Capillary blood from round 1 and 2 was tested for IgG antibodies against SARS-CoV-2 spike proteins with a qualitative chemiluminescent ELISA (spike-only assay). In a subsample of participants (n=82) at round 2, a second assay was performed that measured IgGs reactive to SARS-CoV-2 nucleocapsid protein (nucleocapsid-containing assay). Round 1 biospecimen collections occurred prior to vaccination in all participants. Vaccination status at round 2 was determined via self-report.

**Setting:** The Minneapolis/St. Paul, Minnesota metropolitan area.

**Participants:** HCW age 18-80 years.

**Exposures:** Round 1 recent SARS-CoV-2 infection assessed via a spike-only assay and participant self-report.

**Outcomes:** Round 2 SARS-CoV-2 infection assessed via the nucleocapsid-containing assay. Area under the curve (AUC) was computed to determine the discriminatory ability of round 2 IgG reactivity to nucleocapsid for identification of recent infection determined during round 1

**Results:** Participants had a mean age of 40 (range=23-66) years, 83% were female, 46% reported vaccination prior to the round 2 testing. Round 1 seroprevalence was 9.5%. Among those not recently infected, when comparing vaccinated vs. unvaccinated individuals, elevated levels of spike 1 (p<0.001) and spike 2 (p=0.01) were observed while nucleocapsid levels were not statistically significantly different (p=0.90). Among all participants, nucleocapsid response predicted recent infection with an AUC(95%CI) of 0.93(0.88,0.99). Among individuals vaccinated >10 days prior to antibody testing, the specificity of the nucleocapsid-containing assay was 92% and while the specificity of the spike-only assay was 0%.

**Conclusions and Relevance:** An IgG assay identifying reactivity to nucleocapsid protein is an accurate predictor of natural infection among vaccinated individuals while a spike-only assay performed poorly. In the era of SARS-CoV-2 vaccination, seroprevalence studies monitoring natural infection will require assays that do not rely on spike-protein response alone.

---

Identification of SARS-CoV-2 infection via antibody assays is important for monitoring natural infection rates. Currently authorized antibody tests measure antibody reactivity to spike proteins, yet these develop in response to both infection and vaccination^1,2^. Consequently, ongoing antibody studies will be unable to accurately differentiate prior SARS-CoV-2 infection from vaccination against SARS-CoV-2 in populations with high vaccination coverage.

Current vaccines are not expected to illicit a nucleocapsid response. Consequently, antibody tests that target both spike and nucleocapsid proteins can potentially identify prior infection among vaccinated individuals. The purpose of this study was to compare the accuracy of a nucleocapsid-containing assay vs. a spike protein-only assay in the identification of prior SARS-CoV-2 infection among a sample of healthcare workers (HCW) in the United States.

## METHODS

A sample of healthcare workers located in the Minneapolis/St. Paul, MN, USA, metropolitan area was enrolled^3^ and two rounds of seroprevalence studies were conducted from November 2020–January 2021. A subsample of participants (n=82) for whom excess blood was available for testing with both a spike-only assay and a nucleocapsid-containing assay are presently included. The study was approved by the University of Minnesota Institutional Review Board. All participants provided informed consent.

### Biospecimen Collection and Antibody Assay

Seroprevalence studies occurred in two rounds (Figure 1). Participants collected their first capillary blood specimens between 11/22/2020–12/18/2020 and a second specimen from 12/26/2020–1/08/2021; median time between round 1 and round 2 specimen collections was 28.5 days (range 12–44 days). All round 1 specimens were collected from participants before vaccination. Samples were home-collected using Neoteryx Mitra® 10 µl samplers by volumetric absorption of capillary blood from a finger-stick.

**Figure 1.**
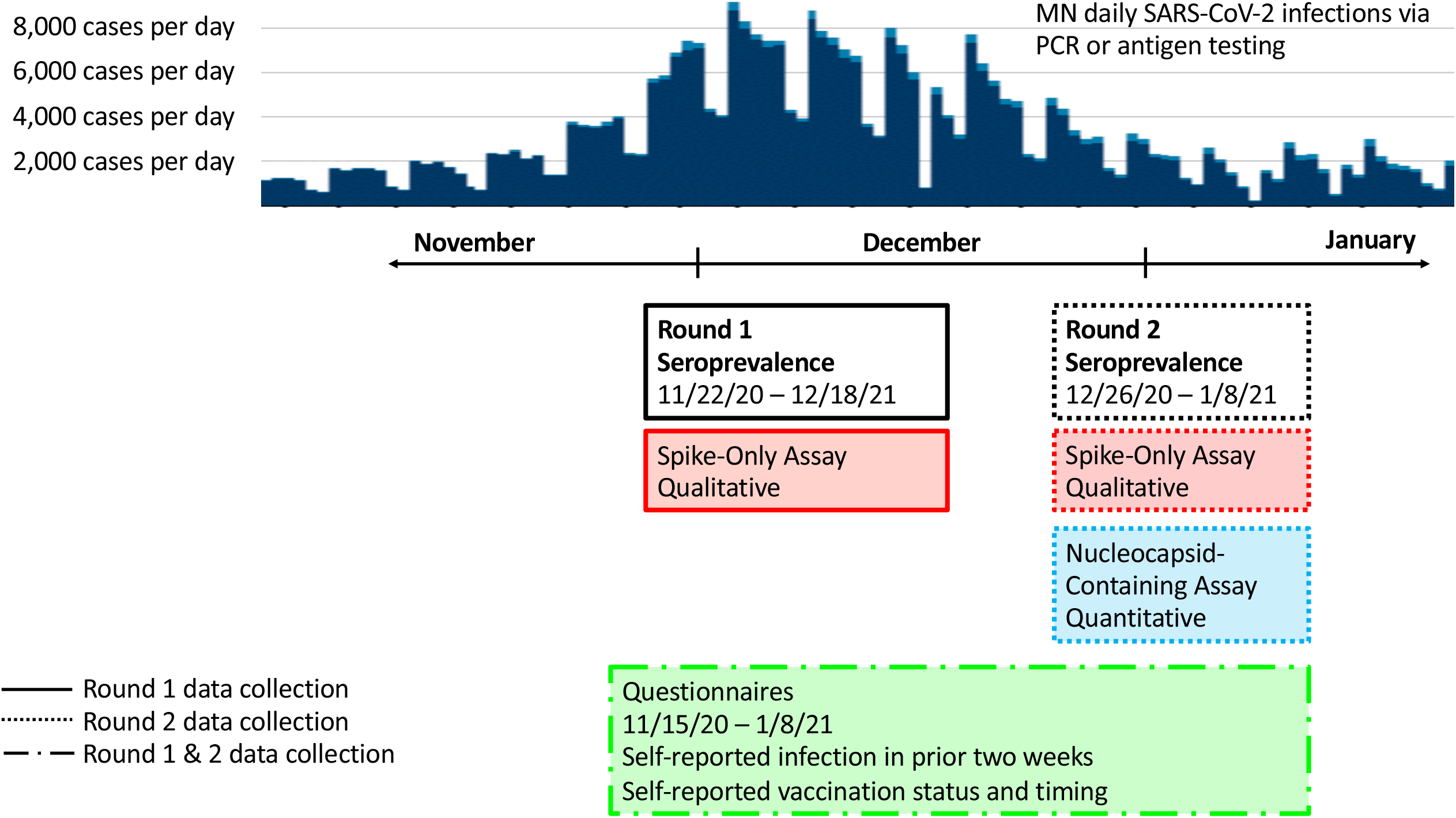
Overview of study design against the backdrop of the Minnesota active infection curve.

Two separate assays were used in this analysis: the Quansys Q-Plex™ SARS-CoV-2 Human IgG (4-Plex), an EUA authorized qualitative chemiluminescent ELISA that concurrently measures Human IgGs reactive to SARS-CoV-2 spike subunit S1 (S1) and spike subunit S2 (S2) (hereafter, ‘spike-only’ assay), and an assay that adds measurement of human IgGs reactive to SARS-CoV-2 nucleocapsid to the spike-only assay (hereafter, ‘nucleocapsid-containing’ assay)

Round 1 samples were tested using the spike-only assay and round 2 samples were tested using both the spike-only assay and the nucleocapsid-containing assay.

Assay details are presented in the supplemental materials.

### Statistical Analysis

‘Recent infection’ with SARS-CoV-2 was defined as a positive result from the round 1 (all round 1 assays were performed prior to vaccination) spike-only assay, or a self-report of recent infection; we consider this to be the definition of ‘true-infection’ (hereafter referred to as ‘infected’ or ‘infection’). Self-reported vaccinations status (yes/no, dose and date) was assessed from questionnaires. Participants were further classified as having been vaccinated 0-7, 7-10, or >10 days prior to collecting capillary blood during round 2.

Kruskal-Wallis tests compared median IgG response to nucleocapsid protein from the nucleocapsid-containing assay at round 2, by vaccination and infection status. Receiver operator characteristic (ROC) curves were constructed and area under the curve (AUC) was computed to determine the discriminatory ability of round 2 IgG reactivity to nucleocapsid for identification of infection. The optimal nucleocapsid cut-point was defined using Youden’s value.^4^ The accuracy of identifying infection from Youden’s optimal nucleocapsid level was then compared to the accuracy of the spike-only assay using round 2 samples.

The accuracy was also estimated for the nucleocapsid-containing assay using the cut-point defined by Youden’s value (described above) in comparison to concurrent round 2 results from the spike-only assay among unvaccinated individuals. In this analysis, infection was defined by results from the concurrent round 2 spike-only assay. These results were then compared against results from the round 2 nucleocapsid-containing assay.

## RESULTS

The mean age of participants was 40 (range=23-66) years and 83% were female. No participants were vaccinated prior to round 1 testing, while 46% of participants reported vaccination prior to the round 2 testing. Among those vaccinated prior to round 2, the mean time between vaccination and antibody testing was 9 days (range=1 to 34). Round one seroprevalence (via the spike-only assay) was 9.5%.

Median values of S1, S2, and nucleocapsid were all statistically significantly elevated among infected vs. uninfected participants (all p-values <0.0001, Figure 2). Among individuals who were not infected, elevated levels of S1 (p=0.0005) and S2 (p=0.01) were observed among vaccinated compared to unvaccinated participants (Figure 2a, 2b) while nucleocapsid levels were not statistically significantly different by vaccination status (p=0.90, Figure 2c).

**Figure 2.**
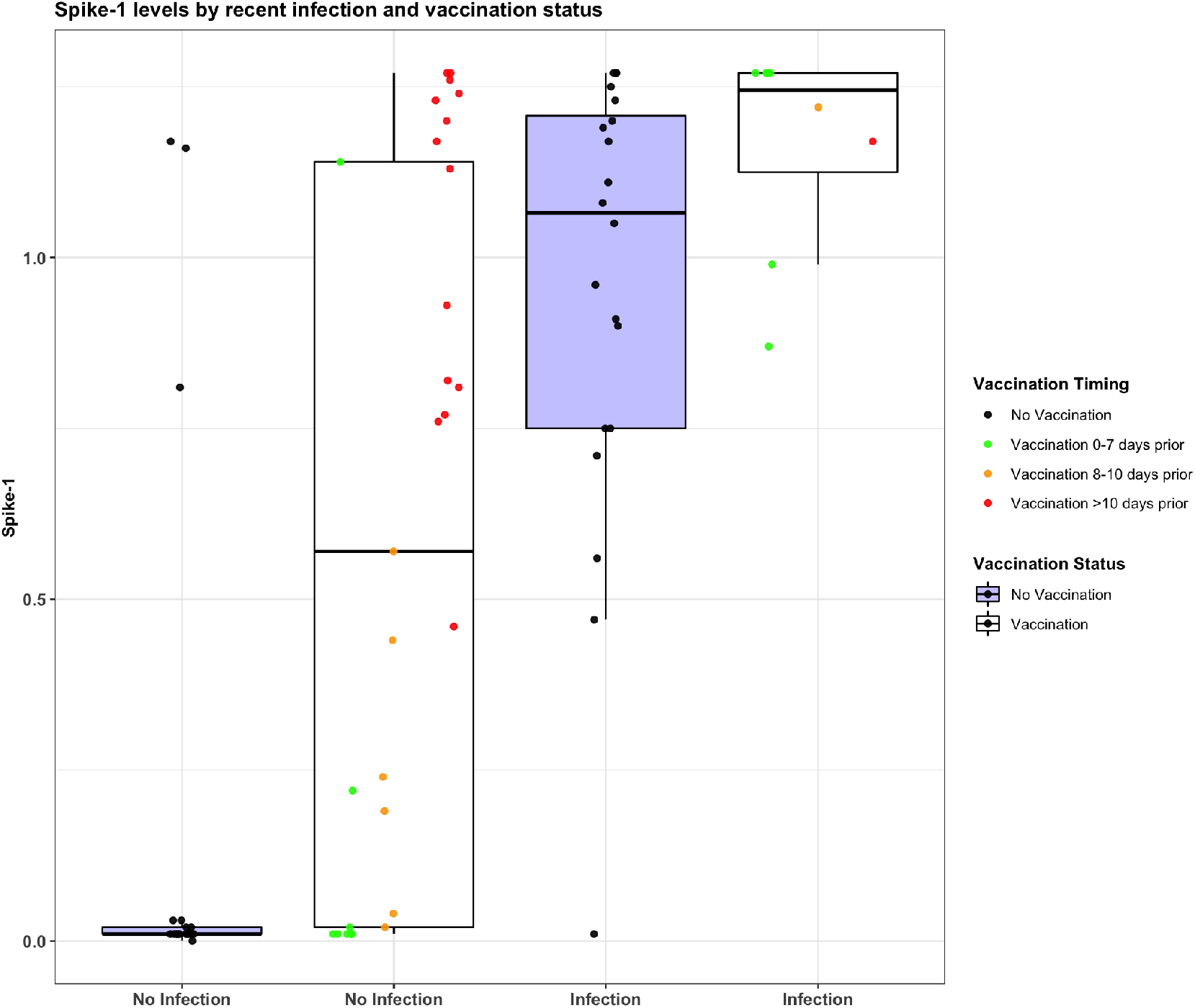
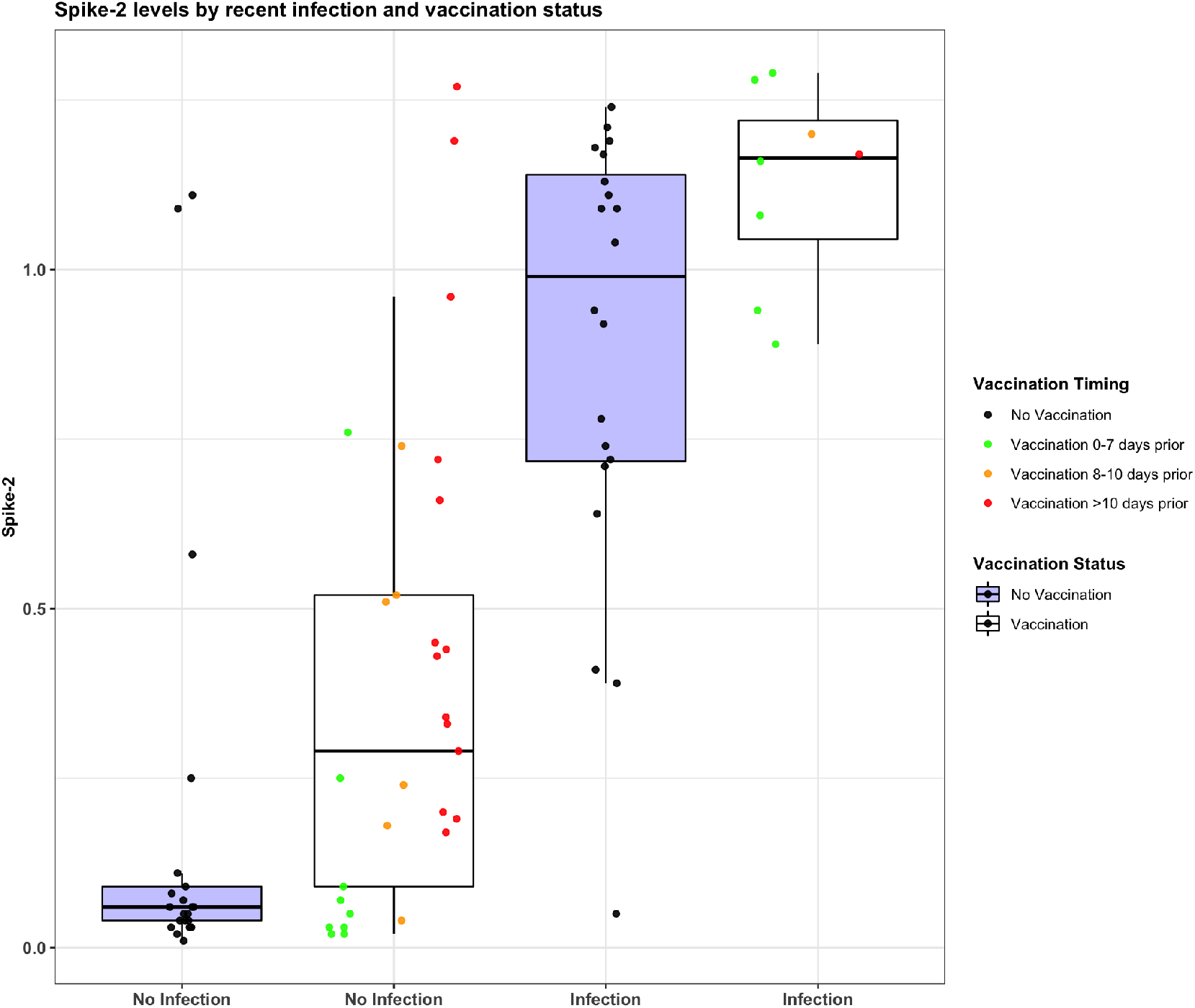
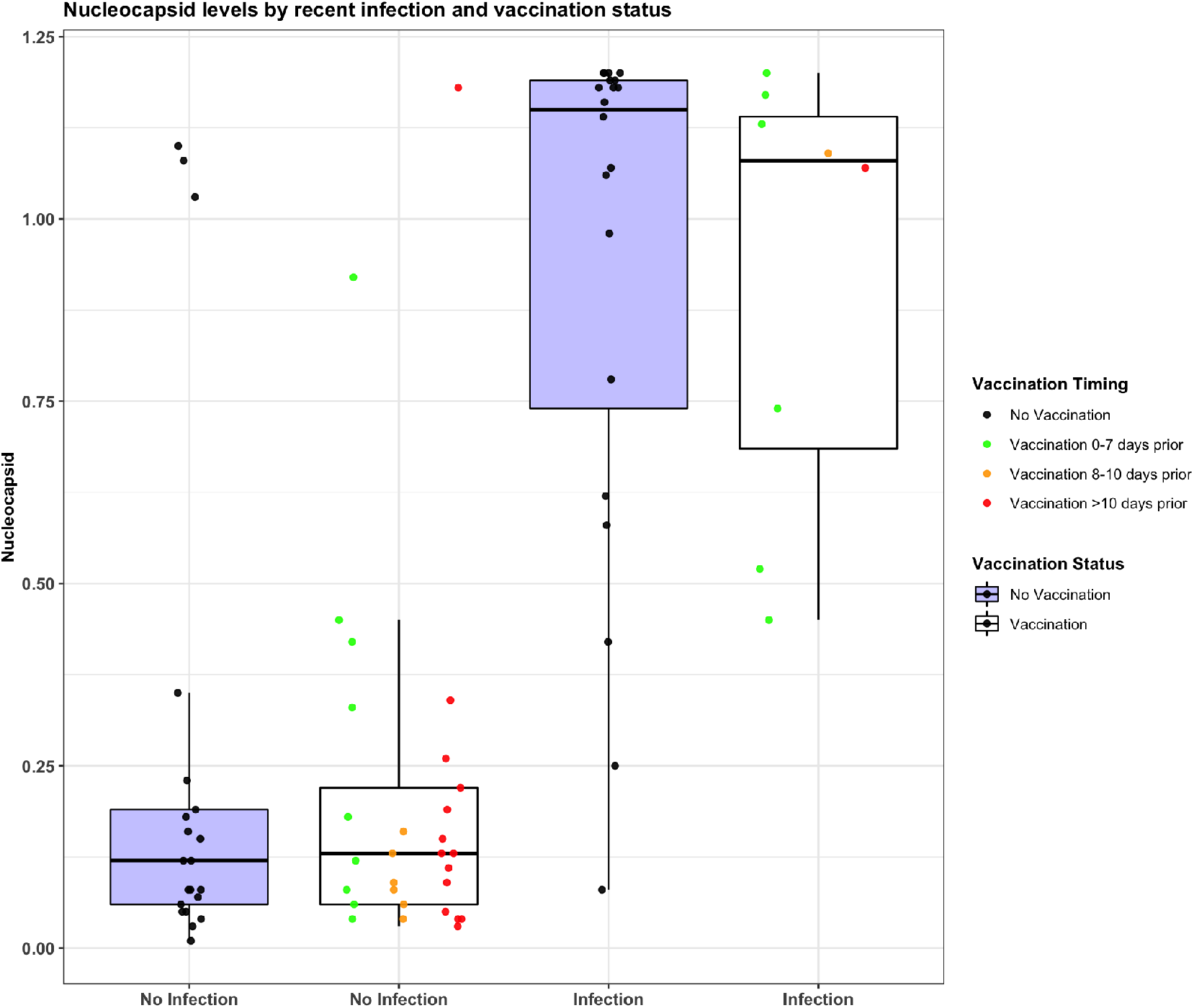
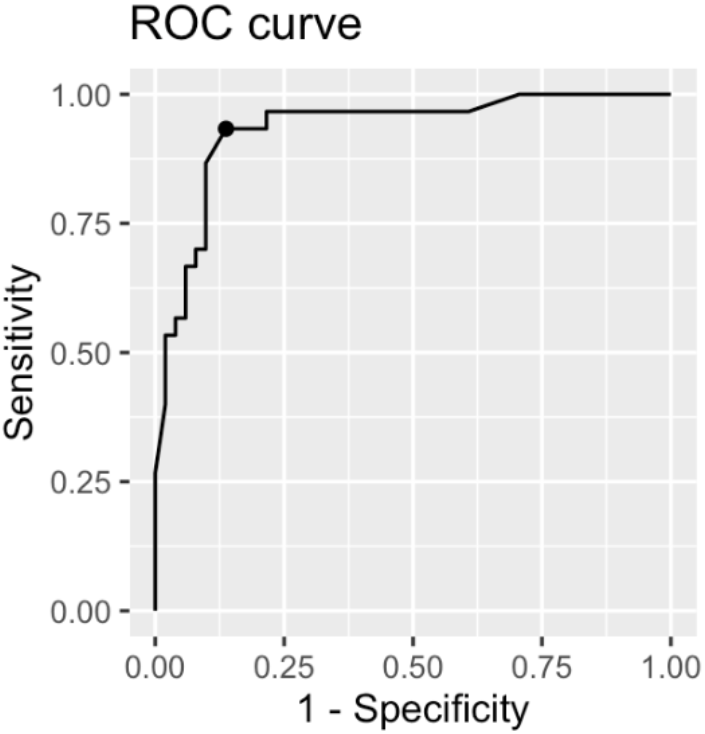
**IgG antibody titers against spike 1 protein (a), and spike 2 protein (b), and nucleocapsid protein (c). Respective p-values for any difference in spike 1, spike 2 or nucleocapsid response by vaccination status among participants without recent infection = 0.0005 (a), 0.01 (b) and 0.90 (c). In panel d, the AUC for the continuous nucleocapsid variable is shown. The area under the curve (AUC) for the continuous nucleocapsid levels = 0.93(0.88,0.99). The optimal nucleocapsid cut-point (Youden’s value) = 0.42 and yields an AUC(95%CI) = 0.89(0.82,0.96).**

Among round 2 samples, nucleocapsid response predicted infection with an AUC(95%CI) of 0.93(0.88,0.99), Figure 2d. Respective AUC(95%CI) values for S1 and S2 were 0.81(0.72,0.90) and 0.89(0.81,0.96). The optimal nucleocapsid cut-point based on Youden’s value was 0.42 and yielded an AUC(95%CI) of 0.89(0.82,0.96) with a sensitivity of 90% and a specificity of 88%. Among the n=37 with vaccination prior to round 2 biospecimen collection, the optimal nucleocapsid cutoff remained at 0.42 with an AUC(95%CI) of 0.95(0.88,1.0), a sensitivity of 90% and specificity of 100%.

Among unvaccinated individuals, sensitivities for the spike-only and nucleocapsid-containing assays were 95% and 85% respectively (Table 1). Among vaccinated individuals both assays had 100% sensitivity. Among unvaccinated individuals the specificity of the spike-only assay was 86% while among individuals vaccinated >10 days before round 2 testing specificity decreased to 0%. In contrast, the specificity of the nucleocapsid assay increases from 86% to 90% among unvaccinated vs. vaccinated individuals. Participants with apparent false positive findings for the nucleocapsid-containing assay (i.e., positive result for the nucleocapsid-containing assay during round 2 and a negative result from the spike-only assay at round 1), the median time between rounds 1 and 2 testing was 33.5 days raising the potential that these individuals were truly infected in the interim.

**Table 1.**
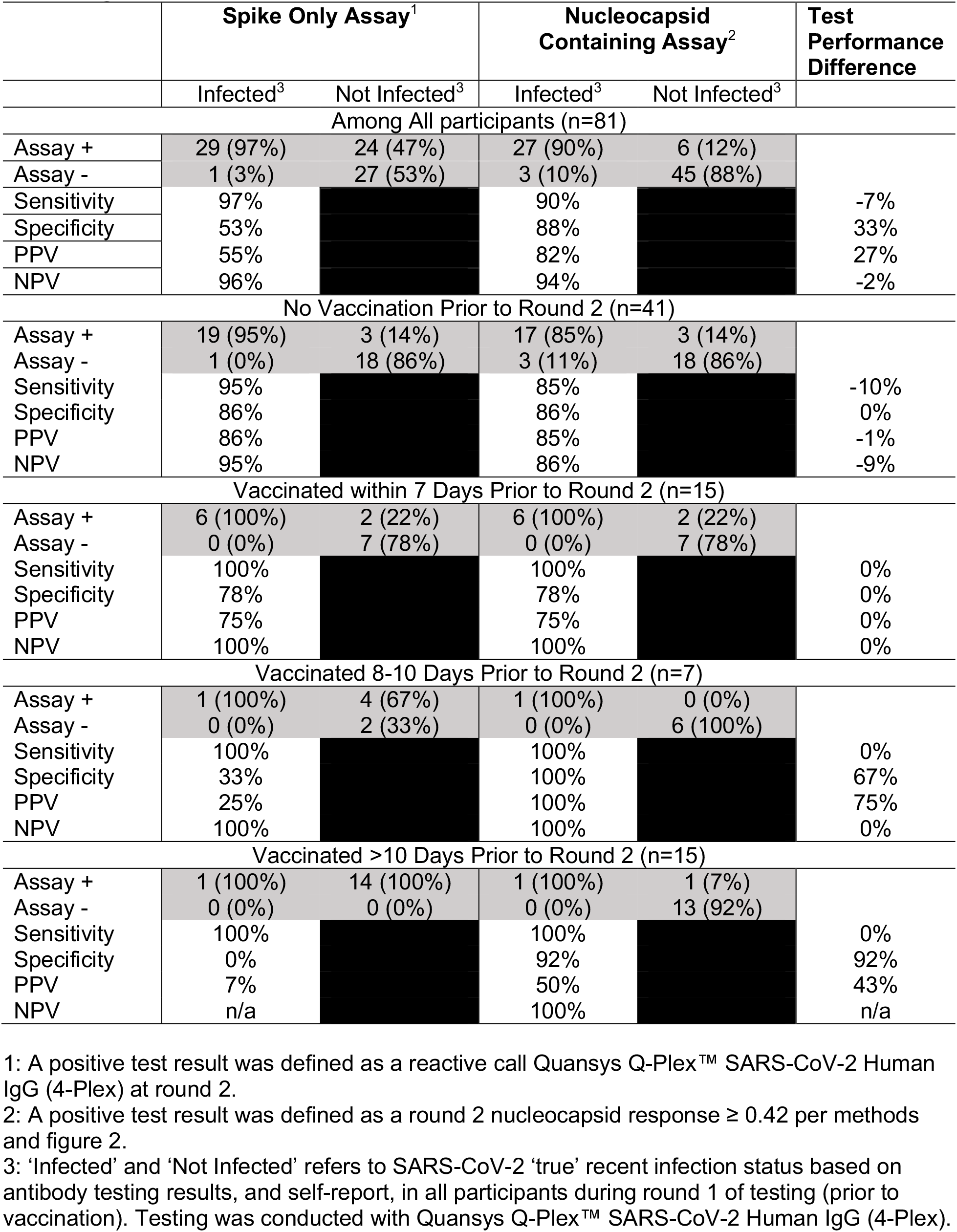
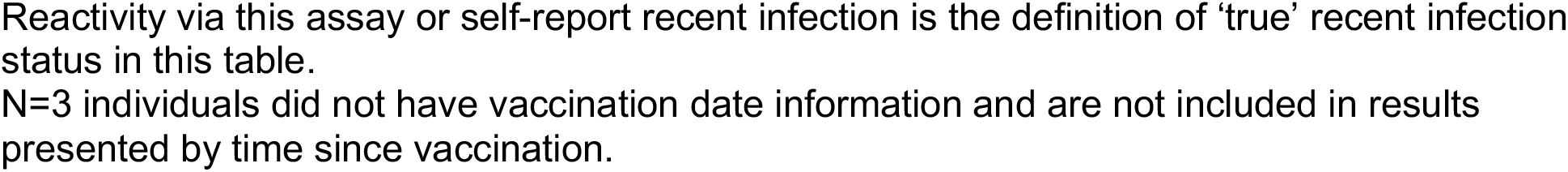
Comparison of SARS-CoV-2 IgG Antibody Test Performance and Predictive Values between a Spike Protein Only Assay and a Nucleocapsid Containing Assay According to Prior Vaccination Status.

Supplemental Table 1 presents test diagnostics results among unvaccinated individuals for the nucleocapsid-containing assay compared to the gold standard spike-only assay test results from the same blood collection time at round 2; this minimizes the potential for false positive findings. Sensitivity and specificity were 90% and 100%, respectively. As expected, when comparing Supplemental Table 1 results to results for the nucleocapsid-containing assay among only the unvaccinated in Table 1, the specificity improves from 86% to 100%, due to fewer false positive findings.

## DISCUSSION

We evaluated the ability of an IgG antibody assay that assesses reactivity to the nucleocapsid protein to identify previous SARS-CoV-2 infection among a population of HCW, with and without infection, and before and after vaccination. This is the first investigation of this nature to our knowledge. We found that in the context of vaccination, nucleocapsid response is highly predictive of infection. Among vaccinated individuals, the nucleocapsid assay had a substantially higher specificity, and positive predictive value, compared to the spike-only assay.

Results in table 1 are limited because infection was determined using an antibody test conducted approximately one month before round two testing, and it is possible that participants appearing as a false-positive in Table 1, were truly infected between round one and round two and developed an antibody response. Therefore, our findings underestimate specificity for the nucleocapsid-containing assay.

An IgG assay identifying reactivity to nucleocapsid protein was an accurate predictor of recent infection among a population of vaccinated HCW. These findings suggest that in the era of SARS-CoV-2 vaccination, seroprevalence studies monitoring natural infection will require assays that capture reactivity to the nucleocapsid protein.

## Data Availability

De-identified data can be made available upon request to Dr. Demmer.

## ON-LINE ONLY SUPPLEMENTAL MATERIALS

Samples were eluted at room temperature (RT, 18-22°C) for 30 minutes on a shaker (500rpm) in 120 µl of a buffer and diluted to the equivalent of 1:100 of venous plasma. The assays are performed at RT with no shaking according to the following steps: (1) 60 minutes incubation of sample/calibrator/control (2) Tris Buffered Saline with Tween (0.05%) (TBST) Wash (3) 30 minutes incubation of biotinylated anti-IgG (4) TBST Wash (5) 20 minute incubation with streptavidin-horseradish peroxidase (6) Wash (7) chemiluminescent substrate (8) Image and analysis using the Q-View™ Imager LS and Q-View™ software. In round 1, a microplate was arrayed with 4 spots in each well: 1) Recombinant SARS-CoV-2 Spike Glycoprotein (S1), comprising amino acids 1-674 of subunit 1, fused with a Sheep Fc-Tag and expressed in HEK293 cells; 2) Recombinant SARS-CoV-2 Spike Glycoprotein (S2), comprising amino acids 685-1211 of subunit 2, fused with a Sheep Fc-Tag and expressed in HEK293 cells; 3) Sheep Fc, a negative control to ensure no cross-reactivity occurs between human IgGs in the sample and the Fc-Tag on the SARS-Cov-2 Spike proteins; 4) Anti-Human IgG is a positive control to ensure that sample was added to the plate and that the procedure was properly followed. Round 2 added a recombinant SARS-CoV-2 nucleocapsid spot, comprising amino acids 1-419, expressed in HEK293 cells.

The spike-only assay used qualitative positive cutoffs for S1 and S2 generated by multiplying a specific correction factor to the ratio of low to high calibrator signal. To determine the semi-quantitative S1, S2 and nucleocapsid ratios for the nucleocapsid-containing assay, the signal (pixel intensity) value for each analyte is divided by the signal of the calibrator high point.

**Supplemental Table 1.** SARS-CoV-2 IgG Nucleocapsid-Containing Antibody Test Performance Among Participants (n=41) Without Prior Vaccination.

|  | Infected<sup>1</sup> | Not Infected<sup>1</sup> |
| --- | --- | --- |
| <sup>2</sup>Assay + | 20 (90%) | 0 (0%) |
| <sup>2</sup>Assay - | 2 (10%) | 19 (19%) |
| Sensitivity | 90% |  |
| Specificity | 100% |  |
| PPV | 100% |  |
| NPV | 90% |  |

1: 'Infected' and 'Not Infected' refers to SARS-CoV-2 recent infection based on antibody testing results among unvaccinated individuals during round 2 of testing. Testing was conducted with Quansys Q-Plex™ SARS-CoV-2 Human IgG (4-Plex) – the spike-only assay. This is the definition of 'true' recent infection status in this table.

## Thank you

We are also profoundly grateful for the study participants who have donated valuable time to advance our understanding about SARS-CoV-2 seroprevalence in healthcare workers.

